# Estimation of the Final Size of the COVID-19 Epidemic in Balochistan, Pakistan

**DOI:** 10.1101/2020.05.22.20110064

**Authors:** Muhammad Arif, Aftab Kakar, Ehsan Larik

**Affiliations:** FELTP

## Abstract

COVID-19 is a new disease that is spreading very fast in Pakistan. Cases have been reported from all the provinces in Pakistan including Balochistan. The first two confirmed cases in Pakistan had travel history from Iran to Pakistan, The some of the initial cases were quarantine at Taftan boarder in Balochistan, hence SIR model is used to predict the magnitude of the disease in Balochistan from May 2020 on wards when lock down and other social distancing measures were loosen up by the government of Balochistan. Our Prediction model shows that about 30,00000 individuals in Balochistan will be infected by 5^th^ of July 2020.Over all 25% of the total population of Balochistan will be affected by this disease with 98% (2940,000) recovery rate by the end of 15^th^ July 2020.

## 1. Introduction

The first case of COVID 19 appeared in Pakistan in the month of Feb 2020, since than the infection has spread all over the country. Cases have been reported from all the four provinces including Balochistan. Since it is a new disease it is highly expected to become endemic in Pakistan very soon. The first case of COVID 19 in Pakistan had travel history by road from Iran to Pakistan, and it had to travel from Balochistan which is situated at the cross road of trade route between Pakistan and Iran.

Since the government of Balochistan has relaxed the lock down measures from May 2020 it will have huge impact on the spread of the disease.Considering this situation the SIR Mathematical modeling can play a key role in predicting the overall impact of COVID-19 in Balochistan.

SIR mathematical model is a compartmental model, it assumes the whole population of Balochistan into three compartments i.e Susceptables (S), Infected (I) and recovered (R□). Every individual in Balochistan will have to pass through all of these compartments.

SIR model operates under the assumption that the total population of Balochistan remains constant and no new births, deaths, in migrants and out migrants are included in the modeling. The model predicts the overall epi-curve by not taking into consideration several factors like age, gender, Lock-down, social distancing, Quarantine and Isolation. The Recovery (R□) compartment assumes a permanent immunity for the infected individuals and also consider the number of individuals that died due to COVID-19 into consideration

## 2. Problem Statement

COVID 19 is a new disease not much is known about it and it is expected become endemic in Pakistan soon.

## 3. Rationale

So far no published study has surveyed the impact of COVID 19 in Balochistan

## 4. Research Questions

Q: When will the Epi-curve for COVI-19 get to its peak in Balochistan?

## 5. Research Methodology

- **Study Design**: Mathematical Prediction Modeling.
- **Study Settings:** District Quetta.
- **Sample Population:** The whole population of Balochistan (i.e 12344408)
- **Data Source:** Daily situation report.
- **Data Analysis:** Vensim Software was used to analyze the data.
- **SIR-Model Assumptions used:**

The model is built on the following set of assumptions, based on the methodology described by fourth-order Runge-Kutta algorithm is expressed as the following differential equations:

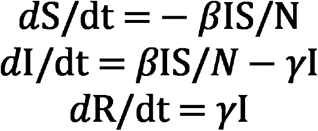

Where:

S: Susceptables
I: Infected
R: Recovered
dt: Rate of
*β*: (Beta) how often a susceptible and infected contact results in a new infection.
*γ:* (Gamma) rate of an infected recovers and moves into recovery phase.

The basic assumption of the SIR model is that the total number of susceptible infected and recovered cases at any given time is equivalent to the test population, so the equations can be represented as:

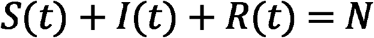

The basic reproduction number (R□), is a ratio between the fraction of individuals susceptible per day (β) and the fraction of recoveries (γ); represented as:

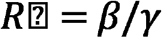

The value of R□ plays a significant role in determining the infectiousness of a certain disease-causing organism. Therefore, the rate of change in infected individuals is directly dependent on the R□, given by:

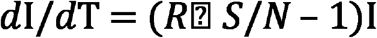

Further assumptions of the SIR model assume that if the R□ is greater than the ratio of total population and the susceptible cases at time zero then it would imply that the outbreak will turn into a full-fledged epidemic.

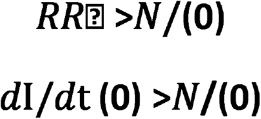

Similarly, if the R□ is less than N/S (0), then it would imply that the outbreak will not cause an Epidemic. Therefore, the R□ plays a crucial role in determining the fate of an epidemic.

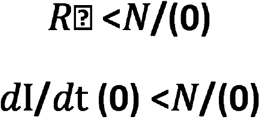

### • Model Parameters

In the case of COVID-19, the value of R□ is highly variable and varies from country to country.Several sources report a range of R□ values, between 1.4 – 3.9, therefore we took an average value of 2.65 for our current analysis. The value of R□ will continue to evolve as the epidemicProgresses throughout the globe. The value of γ was considered based on the average infectious period for COVID-19, so γ=0.14. The value of β was calculated to be 0.378 from equation

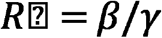

The *S0* was assumed to be 12344408 since the entire population of Balochistan is susceptible to COVID-19, as the disease is new and is spreading across all region.

## 6. Results

**Figure 6.**
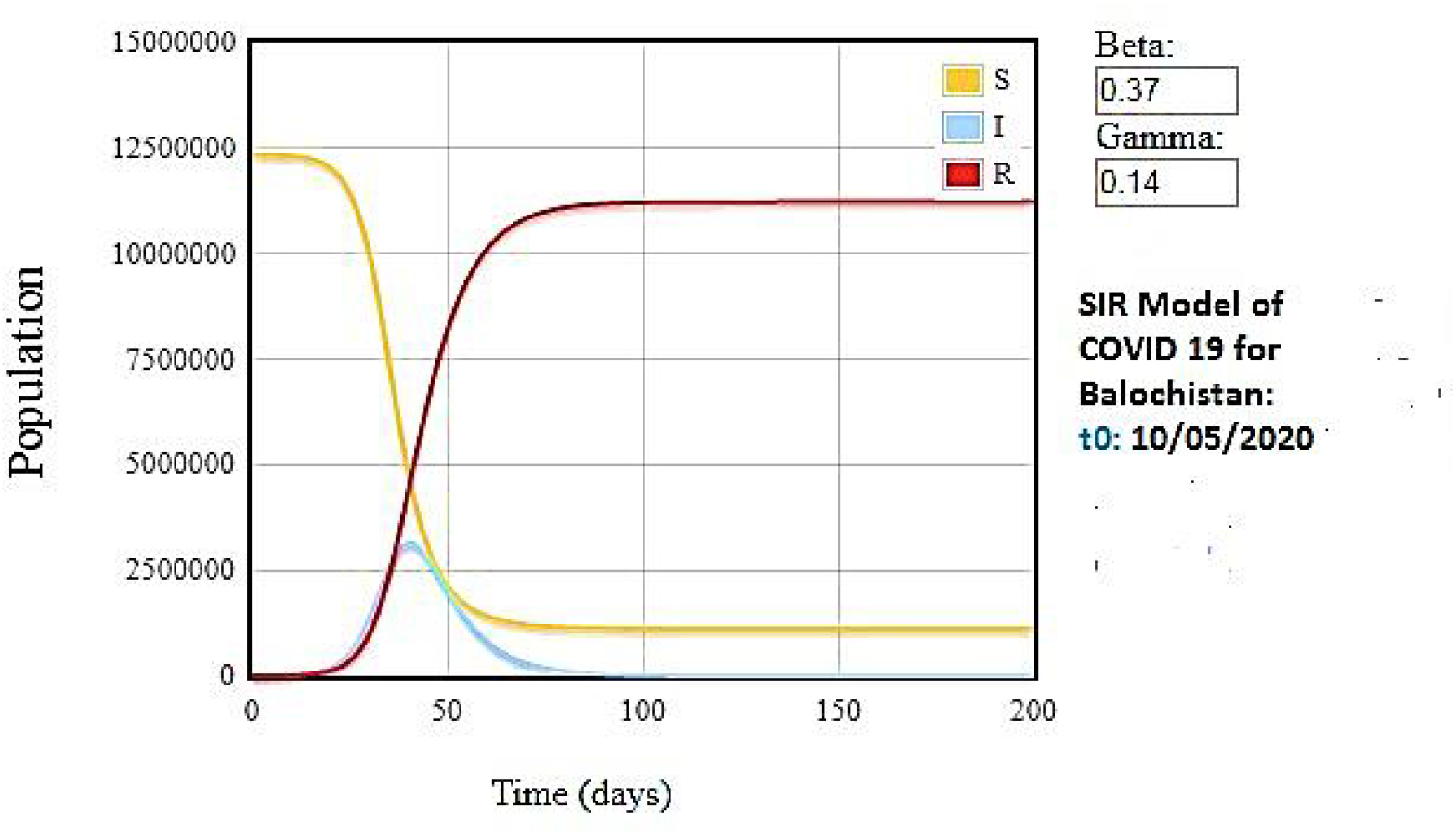
1 The SIR Model of the COVID-19 Epidemic in Balochistan. The simulations suggest that peak infection day will fall on T=45, where I_(45)_ =3000000. The epidemic should have resolved by T=100 where the value of R_(120)_ =2940,000. Susceptible cases (S) shown in Yellow, Infected cases (I) shown in light blue and Recovered cases (R□) shown in red. The x-axis represents the number of days, whereas the y-axis represents the number of cases.

The SIR model for the spread of COVID-19 in Balochistan, under the assumptions mentioned in Model Parameters, indicate that the number of infections will peak on Day 45 (5th July 2020), where 30,00000 individuals could be potentially infected. Over all 25% of the total population of Balochistan will be affected by this disease with 98% (i.e 2940,000) recovery rate by the end of 15th July 2020.

## 7. Discussion

Our study was focused on modelling the COVID-19 epidemic in Balochistan in order to estimate the number of infections, the peak infection day, the rate of increase of infections per day and the resolution of the end-point of the epidemic 1.

The simulation parameters were adjusted according to the population of Balochistan. Our model simulated the conditions where COVID-19 is spreading in a closed population of 12344408 people, without the effect of any extraneous variables such as social distancing, hand washing or travel restrictions. The values of R□=2.65, β=0.378 and γ=0.14 were used.

According to the simulations the peak infection day will occur on 5^th^ July 2020, where 30,00000 persons could get infected with the virus. Previous reports from China have indicated that COVID-19 initially follows an exponential growth pattern ^9^ coupled with asymptomatic carriers ^10^, leads to a rapid increase in the number of infections.

The study starts from the 10^th^ May 2020 using the situation report and prior to 200 days the peak is predicted to have reached with 30,00000 individuals infected that is almost 25% of the population will be infected till 5^th^ of July 2020.

However, the major concern for Balochistan would be the healthcare system which would not be able to cope with the overwhelming number of patients if the trajectory remains the same. Studies place the mortality rate of COVID-19 at 2.3%, severe cases at 14% and critical cases at 5%^12,13^, which would imply that potentially 60,000 could die; 420,000 cases could become severe and 150,000 could become critical during the aftermath of epidemic in Balochistan. Therefore, there is an urgent need to implement effective measures to curb the rise in COVID-19 infections in Balochistan, otherwise it could lead to drastic consequences.

## 8. Recommendations

- Mass media awareness for COVID-19.
- Ensuring the SOPs of social distancing and face mask.
- Promoting and ensuring hand hygiene practices.

## Data Availability

primary data

